# Tixagevimab/Cilgavimab for Prevention of COVID-19 during the Omicron Surge: Retrospective Analysis of National VA Electronic Data

**DOI:** 10.1101/2022.05.28.22275716

**Authors:** Yinong Young-Xu, Lauren Epstein, Vincent C Marconi, Victoria Davey, Gabrielle Zwain, Jeremy Smith, Caroline Korves, Fran Cunningham, Robert Bonomo, Adit A Ginde

## Abstract

**Background:** Little is known regarding the effectiveness of tixagevimab/cilgavimab in preventing SARS-CoV-2 infection in this population, particularly after the emergence of the Omicron variant.

**Objective:** To determine the effectiveness of tixagevimab/cilgavimab for prevention of SARS-CoV-2 infection and severe disease among immunocompromised patients.

**Design:** Retrospective cohort study with propensity matching and difference-in-difference analyses.

**Setting:** U.S. Department of Veterans Affairs (VA) healthcare system.

**Participants:** Veterans age ≥18 years as of January 1, 2022, receiving VA healthcare. We compared a cohort of 1,848 patients treated with at least one dose of intramuscular tixagevimab/cilgavimab to matched controls selected from 251,756 patients who were on immunocompromised or otherwise at high risk for COVID-19. Patients were followed through April 30, 2022, or until death, whichever occurred earlier.

**Main Outcomes:** Composite of SARS-CoV-2 infection, COVID-19-related hospitalization, and all-cause mortality. We used cox proportional hazards modelling to estimate the hazard ratios (HR) and 95% CI for the association between receipt of tixagevimab/cilgavimab and outcomes.

**Results:** Most (69%) tixagevimab/cilgavimab recipients were ≥65 years old, 92% were identified as immunocompromised in electronic data, and 73% had ≥3 mRNA vaccine doses or two doses of Ad26.COV2. Compared to propensity-matched controls, tixagevimab/cilgavimab-treated patients had a lower incidence of the composite COVID-19 outcome (17/1733 [1.0%] vs 206/6354 [3.2%]; HR 0.31; 95%CI, 0.18-0.53), and individually SARS-CoV-2 infection (HR 0.34; 95%CI, 0.13-0.87), COVID-19 hospitalization (HR 0.13; 95%CI, 0.02-0.99), and all-cause mortality (HR 0.36; 95%CI, 0.18-0.73).

**Limitations:** Confounding by indication and immortal time bias.

**Conclusions:** Using national real-world data from predominantly vaccinated, immunocompromised Veterans, administration of tixagevimab/cilgavimab was associated with lower rates of SARS-CoV-2 infection, COVID-19 hospitalization, and all-cause mortality during the Omicron surge.

## INTRODUCTION

Immunocompromised patients are at high risk for morbidity and mortality related to COVID-19.^1^ While vaccines have helped to prevent the spread of SARS-CoV-2 and decrease the risk of severe disease in the general population, immunocompromised patients remain at higher risk for breakthrough infections and persistent viral replication.^2,3,4,5^

The PROVENT study, a Phase 3, multicenter, randomized, placebo-controlled trial, demonstrated a single dose of intramuscular tixagevimab/cilgavimab (Evusheld, AstraZeneca) significantly reduced the incidence of symptomatic SARS-CoV-2 infection by 76.7% after 90 days in a broad population of adults with an increased risk of inadequate response to vaccination and/or increased risk of exposure to SARS-CoV-2.^6^ Based on these findings, on December 8, 2021, the US Food and Drug Administration (FDA) granted an emergency use authorization (EUA) of tixagevimab/cilgavimab as pre-exposure prophylaxis for moderate to severe immune compromised individuals or for whom vaccination with any available COVID-19 vaccine is not recommended due to a history of severe adverse reaction.^7^ The PROVENT trial also included those with chronic health conditions that could put individuals at elevated risk for complications owing to COVID-19.

Importantly, questions remain regarding effectiveness of tixagevimab/cilgavimab for the prevention of COVID-19. Only a small proportion (11%) of participants in the PROVENT trial were immunocompromised (i.e., receipt of immunosuppressive therapy, have immunosuppressive disease or cancer), and treatment effectiveness in this crucial subgroup could not be estimated in the trial.

Furthermore, all participants in the PROVENT trial were unvaccinated at the time of trial entry; therefore, indications for tixagevimab/cilgavimab among vaccinated persons is unknown. Finally, follow-up of participants in the PROVENT trial ended in September 2021; therefore, an analysis regarding real-world effectiveness is needed for tixagevimab/cilgavimab among vaccinated immunocompromised patients after the emergence of the Omicron variant (December 2021 in the US).^8^

Our objective was to assess the effectiveness of tixagevimab/cilgavimab for prevention of COVID-19 during the Omicron surge using electronic data from the U.S. Department of Veterans Affairs (VA), the largest integrated health care system in the US., the largest integrated healthcare system in the US. Using propensity score matching and Difference-in-Difference (DiD) approaches, we estimated the real-world effectiveness of tixagevimab/cilgavimab among immunocompromised Veterans for the prevention of SARS-CoV-2 infection, COVID-19 related hospitalization, and all-cause mortality.

## METHODS

### Study Setting and Data Sources

The VA provides care to nearly 9 million Veterans at 171 medical centers and 1112 outpatient clinics across the US. The first dose of tixagevimab/cilgavimab was given at VA on January 13, 2022. We analyzed electronic health records (EHR) using the VA Corporate Data Warehouse (CDW), which contains patient-level information on all patient encounters in VA medical facilities, including treatments, prescriptions, vaccinations, laboratory results, healthcare utilization, and vital status.^9,^10 We identified tixagevimab/cilgavimab use through the VA Pharmacy Benefits Management (PBM) EUA prescription dashboard, which captures and links records of recipients, date, and dosage of tixagevimab/cilgavimab administered in medical facilities across VA.^11^

This study was approved by the institutional review board of the VA Medical Center in White River Junction, Vermont, and was granted a waiver of informed consent because the study was deemed minimal risk and consent impractical to acquire. This study followed the Strengthening the Reporting of Observational Studies in Epidemiology (<u>STROBE</u>) reporting guideline.

### Study population and outcomes

We included Veterans who were ≥18 years (as of January 1, 2022) and received VA healthcare through April 30, 2022 or until death, whichever occurred earlier, and who fulfilled study period and other inclusion criteria. Tixagevimab/cilgavimab (150 mg/150 mg) was first administered in the VA on January 13, 2022. On February 24, 2022, in response to concerns regarding effectiveness of the tixagevimab/cilgavimab against the Omicron variant, the FDA revised the EUA to increase the initial dose tixagevimab/cilgavimab to 300 mg/300 mg; patients who received the previously authorized (lower) dose were advised to receive an additional dose.^12^ In our current analysis, we included any patient who received at least one dose of tixagevimab/cilgavimab during the observation period in the treatment arm. Controls were immunocompromised or other high-risk patients who did not receive tixagevimab/cilgavimab. To address immortal time bias control patients were assigned pseudo-administration dates to match the real treatment dates of the tixagevimab/cilgavimab patient cohort. These dates, real and imitated, served as the index date of follow-up. The study period was then divided into phases. We looked back over a maximum of two years before the date of treatment to assess baseline characteristics, with a follow-up period from date of receiving tixagevimab/cilgavimab through April 30, 2022, or until death, whichever occurred earlier.

Characteristics measured during the baseline period included demographics, significant comorbidities, and healthcare utilization. We used VA-assigned priority group for healthcare to serve as a surrogate measure for socioeconomic status.^13^ Information regarding comorbidities was abstracted from diagnosis codes recorded in VA electronic data for healthcare encounters during any VA visit in the two years before the index date; significant comorbidities were defined according to an adaptation of Deyo-Charlson comorbidity index (DCCI).^14^ We defined immunocompromised status based on 1) whether the patient received an immunosuppressive medication during the 30 days before the index date (Appendix I) or 2) the presence of at least one qualifying immunocompromising condition, based on ICD-10 code listed in Appendix II, during the two years before index date.^15^ We defined severely immunocompromised as those who had a solid organ transplant or received anti-rejection medication for transplant or chemotherapy for cancer treatment in the prior month.

The primary outcome was the composite of 1) SARS-CoV-2 infections confirmed by the presence of SARS-CoV-2 virus detected by reverse-transcriptase-polymerase-chain-reaction (RT-PCR) or antigen testing; 2) COVID-19 hospitalization, defined as having both an admission and discharge diagnosis for COVID-19 from a hospital or within 30 days of positive SARS-CoV-2 RT-PCR result or antigen test; 3) all-cause mortality, defined as having a date of death (DoD) during the follow-up. Clinical visits with urinary tract infection (UTI) as the primary discharge diagnosis (UTI, ICD-9: 599) were added as a fourth outcome in falsification test as for a negative control.^16^

### Statistical Analysis

#### Propensity Score Matching

We used propensity score models to account for observable baseline differences between patients who received tixagevimab/cilgavimab and controls. All covariates in the propensity score (Appendix III) were measured before the initiation of tixagevimab/cilgavimab to avoid adjustment for potential mediators. Indicator variables were generated to capture missing or unknown values for any of the matching criteria to retain patients in the study. Propensity score matching was performed using greedy nearest neighbor matching with caliper of 0.2 and ratio of 1:4 with replacement.^17^ In order to assess the robustness of the propensity match, we calculated the standardized mean difference (SMD); a successful match was estimated when at least 90% of the covariates included in the propensity score model had Standardized Mean Difference (SMD) of ≤10.^18^

To address immortal time bias,^19^ we generated a pseudo-tixagevimab/cilgavimab administration date for each control that follows the same distribution as those actual administration dates for recipients of tixagevimab/cilgavimab.^20^ In the final model, we matched patients who received tixagevimab/cilgavimab and eligible controls based on the date (or pseudo-date) and the facility where tixagevimab/cilgavimab was administered. We excluded any patients who were diagnosed with SARS-CoV-2 infection via a positive RT-PCR result or antigen testing within 3 months of the date or pseudo-date of tixagevimab/cilgavimab administration to optimize focus on new infections. We used Cox proportional hazards regression to compare patients who received tixagevimab/cilgavimab and their matched controls.

#### Difference-in-Difference Analysis

In addition to the propensity score model, we used a difference-in-difference (DiD) analysis to assess outcomes. The DiD analysis is a quasi-experimental method used to estimate the causal effect of an intervention.^21^ We calculated a person-time denominator for patients who received tixagevimab/cilgavimab and controls by tallying the number of days those patients were enrolled for an extended study period (September 1, 2021 through April 30, 2022). We calculated a numerator as total number of outcomes (including multiple outcomes for a single patient) per period. Outcome rates were then calculated for treated and controls during the baseline (last 4 months of 2021) and observation period. To simplify, we calculated events by calendar month. This was only employed to show the background rates of events during this extended period for the unmatched study population.

After propensity-score matching, we adjusted for residual confounding using the prior event rate ratio (PERR) approach.^22^ This method, like the difference-in-differences method used in econometrics, accounts for two distinct time periods, time before the intervention (e.g., tixagevimab/cilgavimab administration date and pseudo-tixagevimab/cilgavimab administration date) and time after the intervention. For each cohort, the rates of each outcome were calculated and compared before and after the intervention within the extended study period. To assess the impact of the intervention, the relative rate of the post-treatment period was divided by the relative rate of the pre-treatment period.

To apply the PERR method, we first computed the incidence rate ratio (RR) for each study outcome in the observation period (RR_o_) and then again in the baseline period (RR_b_). The RR is the rate of the outcome among tixagevimab/cilgavimab recipients divided by the rate of the outcome in the control arm. Next, we computed the PERR per the following formula PERR=(RRo / RR_b_), and, finally, the relative effectiveness of tixagevimab/cilgavimab to SPM (rE) is defined as (1-PERR) x 100%.

Analyses were performed with Stata 17 software (StataCorp), and SAS software, version 8.2 (SAS Institute).

## RESULTS

### Study Population

We identified 1,848 recipients of tixagevimab/cilgavimab and 251,756 control patients who were immunocompromised or otherwise at high risk for COVID-19 (**Figure 1**). After propensity-score matching 1,733 remained in the treatment group and 6,354 in the control group, which were well balanced across baseline characteristics (**Table 1**). Among the treated, 1,579 (91%) were male, 277 (16%) were Black, 76 (4%) were Hispanic, and 1250 (72%) were non-Hispanic White. Most (1,187 [69%]) of the tixagevimab/cilgavimab recipients were ≥ 65 years old and 1,238 (71%) lived in urban areas.^23^ We identified 1,595 (92%) tixagevimab/cilgavimab recipients as immunocompromised in the electronic data (by immunosuppressants use and diagnosis codes in Appendix Tables I, II). The propensity-matched control group had the same proportion of immunocompromised patients (5,863 [92%]). Additional common comorbidities in treated patients included 1,029 (59%) hypertension, 612 (35%) dyslipidemia, 597 (34%) cancer, and the majority were overweight (BMI [kg/m^2^] ≥ 25.0 and <30, 674 [39%]) or obese (BMI 30.0 or higher, 632 [36%]). Most tixagevimab/cilgavimab recipients had received 2 doses of mRNA vaccines or 1 dose of Ad26.COV2 (Janssen) (22%) or ≥ 3 vaccine doses or 2 doses of Ad26.COV2 (73%). Only 88 (5%) tixagevimab/cilgavimab recipients did not have any record of COVID-19 vaccination, compared to 67,753 (27%) among unmatched control patients.

**Table 1.**
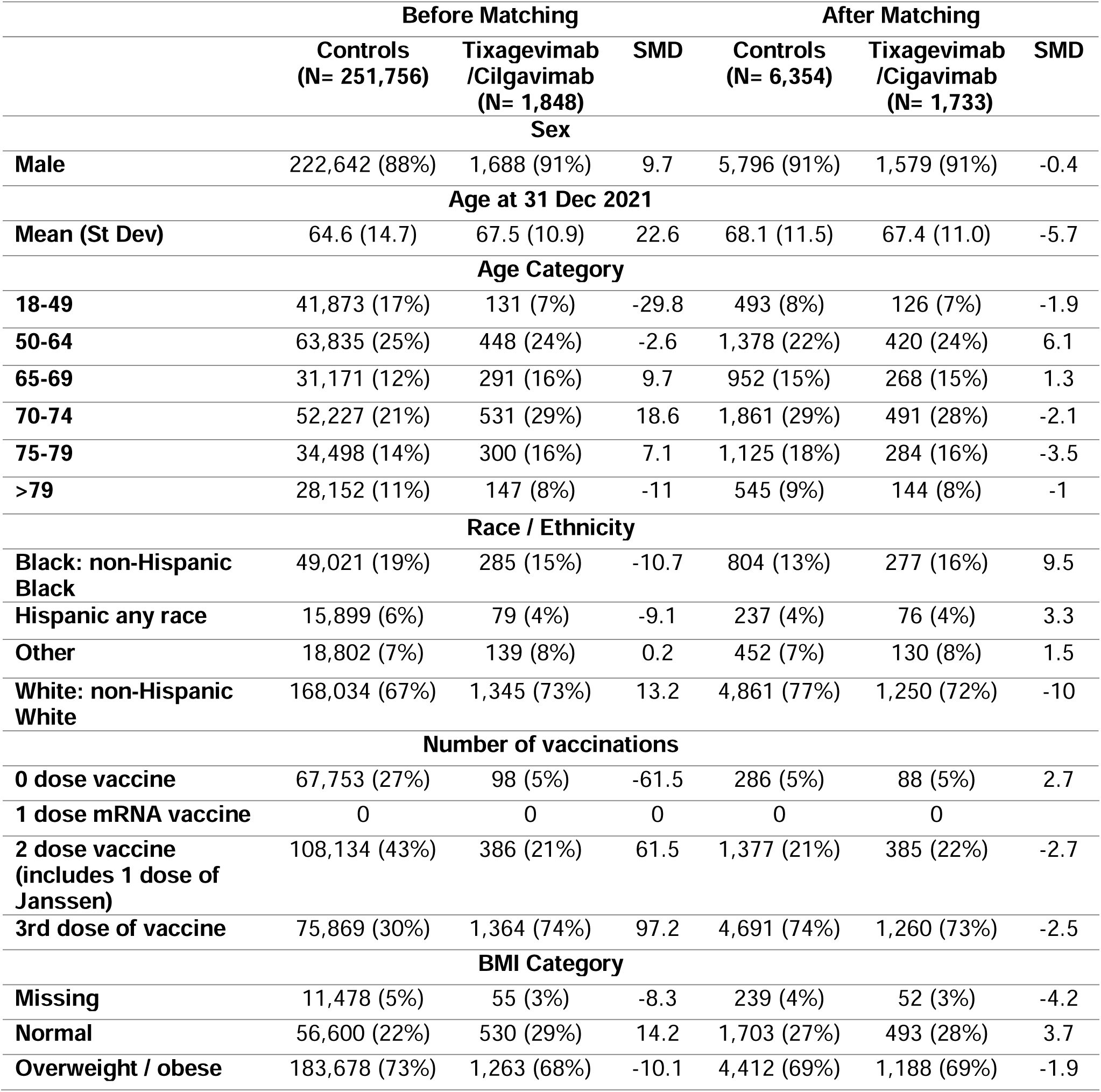
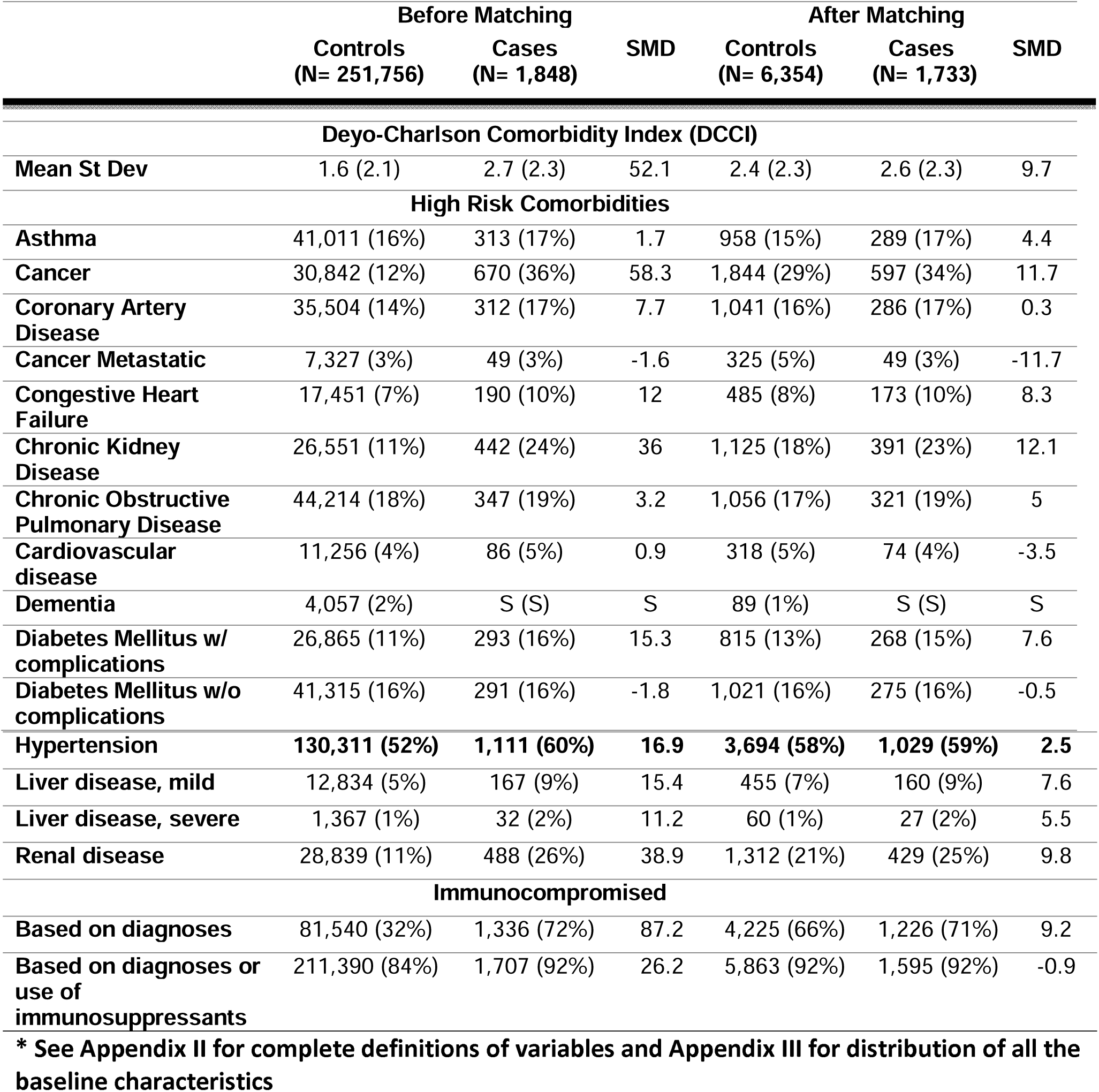
Selected* Baseline Characteristics.

|  | Before Matching |  |  | After Matching |  |  |
| --- | --- | --- | --- | --- | --- | --- |
|  | Controls<br>(N= 251,756) | Tixagevimab<br>/Cilgavimab<br>(N= 1,848) | SMD | Controls<br>(N= 6,354) | Tixagevimab<br>/Cilgavimab<br>(N= 1,733) | SMD |
| **Sex** |  |  |  |  |  |  |
| **Male** | 222,642 (88%) | 1,688 (91%) | 9.7 | 5,796 (91%) | 1,579 (91%) | -0.4 |
| **Age at 31 Dec 2021** |  |  |  |  |  |  |
| **Mean (St Dev)** | 64.6 (14.7) | 67.5 (10.9) | 22.6 | 68.1 (11.5) | 67.4 (11.0) | -5.7 |
| **Age Category** |  |  |  |  |  |  |
| **18-49** | 41,873 (17%) | 131 (7%) | -29.8 | 493 (8%) | 126 (7%) | -1.9 |
| **50-64** | 63,835 (25%) | 448 (24%) | -2.6 | 1,378 (22%) | 420 (24%) | 6.1 |
| **65-69** | 31,171 (12%) | 291 (16%) | 9.7 | 952 (15%) | 268 (15%) | 1.3 |
| **70-74** | 52,227 (21%) | 531 (29%) | 18.6 | 1,861 (29%) | 491 (28%) | -2.1 |
| **75-79** | 34,498 (14%) | 300 (16%) | 7.1 | 1,125 (18%) | 284 (16%) | -3.5 |
| **>79** | 28,152 (11%) | 147 (8%) | -11 | 545 (9%) | 144 (8%) | -1 |
| **Race / Ethnicity** |  |  |  |  |  |  |
| **Black: non-Hispanic Black** | 49,021 (19%) | 285 (15%) | -10.7 | 804 (13%) | 277 (16%) | 9.5 |
| **Hispanic any race** | 15,899 (6%) | 79 (4%) | -9.1 | 237 (4%) | 76 (4%) | 3.3 |
| **Other** | 18,802 (7%) | 139 (8%) | 0.2 | 452 (7%) | 130 (8%) | 1.5 |
| **White: non-Hispanic White** | 168,034 (67%) | 1,345 (73%) | 13.2 | 4,861 (77%) | 1,250 (72%) | -10 |
| **Number of vaccinations** |  |  |  |  |  |  |
| **0 dose vaccine** | 67,753 (27%) | 98 (5%) | -61.5 | 286 (5%) | 88 (5%) | 2.7 |
| **1 dose mRNA vaccine** | 0 | 0 | 0 | 0 | 0 |  |
| **2 dose vaccine**<br>**(includes 1 dose of Janssen)** | 108,134 (43%) | 386 (21%) | 61.5 | 1,377 (21%) | 385 (22%) | -2.7 |
| **3rd dose of vaccine** | 75,869 (30%) | 1,364 (74%) | 97.2 | 4,691 (74%) | 1,260 (73%) | -2.5 |
| **BMI Category** |  |  |  |  |  |  |
| **Missing** | 11,478 (5%) | 55 (3%) | -8.3 | 239 (4%) | 52 (3%) | -4.2 |
| **Normal** | 56,600 (22%) | 530 (29%) | 14.2 | 1,703 (27%) | 493 (28%) | 3.7 |
| **Overweight / obese** | 183,678 (73%) | 1,263 (68%) | -10.1 | 4,412 (69%) | 1,188 (69%) | -1.9 |
|  | Controls<br>(N= 251,756) | Cases<br>(N= 1,848) | SMD | Controls<br>(N= 6,354) | Cases<br>(N= 1,733) | SMD |
| **Deyo-Charlson Comorbidity Index (DCCI)** |  |  |  |  |  |  |
| Mean St Dev | 1.6 (2.1) | 2.7 (2.3) | 52.1 | 2.4 (2.3) | 2.6 (2.3) | 9.7 |
| **High Risk Comorbidities** |  |  |  |  |  |  |
| Asthma | 41,011 (16%) | 313 (17%) | 1.7 | 958 (15%) | 289 (17%) | 4.4 |
| Cancer | 30,842 (12%) | 670 (36%) | 58.3 | 1,844 (29%) | 597 (34%) | 11.7 |
| Coronary Artery Disease | 35,504 (14%) | 312 (17%) | 7.7 | 1,041 (16%) | 286 (17%) | 0.3 |
| Cancer Metastatic | 7,327 (3%) | 49 (3%) | -1.6 | 325 (5%) | 49 (3%) | -11.7 |
| Congestive Heart Failure | 17,451 (7%) | 190 (10%) | 12 | 485 (8%) | 173 (10%) | 8.3 |
| Chronic Kidney Disease | 26,551 (11%) | 442 (24%) | 36 | 1,125 (18%) | 391 (23%) | 12.1 |
| Chronic Obstructive Pulmonary Disease | 44,214 (18%) | 347 (19%) | 3.2 | 1,056 (17%) | 321 (19%) | 5 |
| Cardiovascular disease | 11,256 (4%) | 86 (5%) | 0.9 | 318 (5%) | 74 (4%) | -3.5 |
| Dementia | 4,057 (2%) | S (S) | S | 89 (1%) | S (S) | S |
| Diabetes Mellitus w/ complications | 26,865 (11%) | 293 (16%) | 15.3 | 815 (13%) | 268 (15%) | 7.6 |
| Diabetes Mellitus w/o complications | 41,315 (16%) | 291 (16%) | -1.8 | 1,021 (16%) | 275 (16%) | -0.5 |
| Hypertension | **130,311 (52%)** | **1,111 (60%)** | **16.9** | **3,694 (58%)** | **1,029 (59%)** | **2.5** |
| Liver disease, mild | 12,834 (5%) | 167 (9%) | 15.4 | 455 (7%) | 160 (9%) | 7.6 |
| Liver disease, severe | 1,367 (1%) | 32 (2%) | 11.2 | 60 (1%) | 27 (2%) | 5.5 |
| Renal disease | 28,839 (11%) | 488 (26%) | 38.9 | 1,312 (21%) | 429 (25%) | 9.8 |
| **Immunocompromised** |  |  |  |  |  |  |
| Based on diagnoses | 81,540 (32%) | 1,336 (72%) | 87.2 | 4,225 (66%) | 1,226 (71%) | 9.2 |
| Based on diagnoses or use of immunosuppressants | 211,390 (84%) | 1,707 (92%) | 26.2 | 5,863 (92%) | 1,595 (92%) | -0.9 |

\* See Appendix II for complete definitions of variables and Appendix III for distribution of all the baseline characteristics

**Figure 1.**
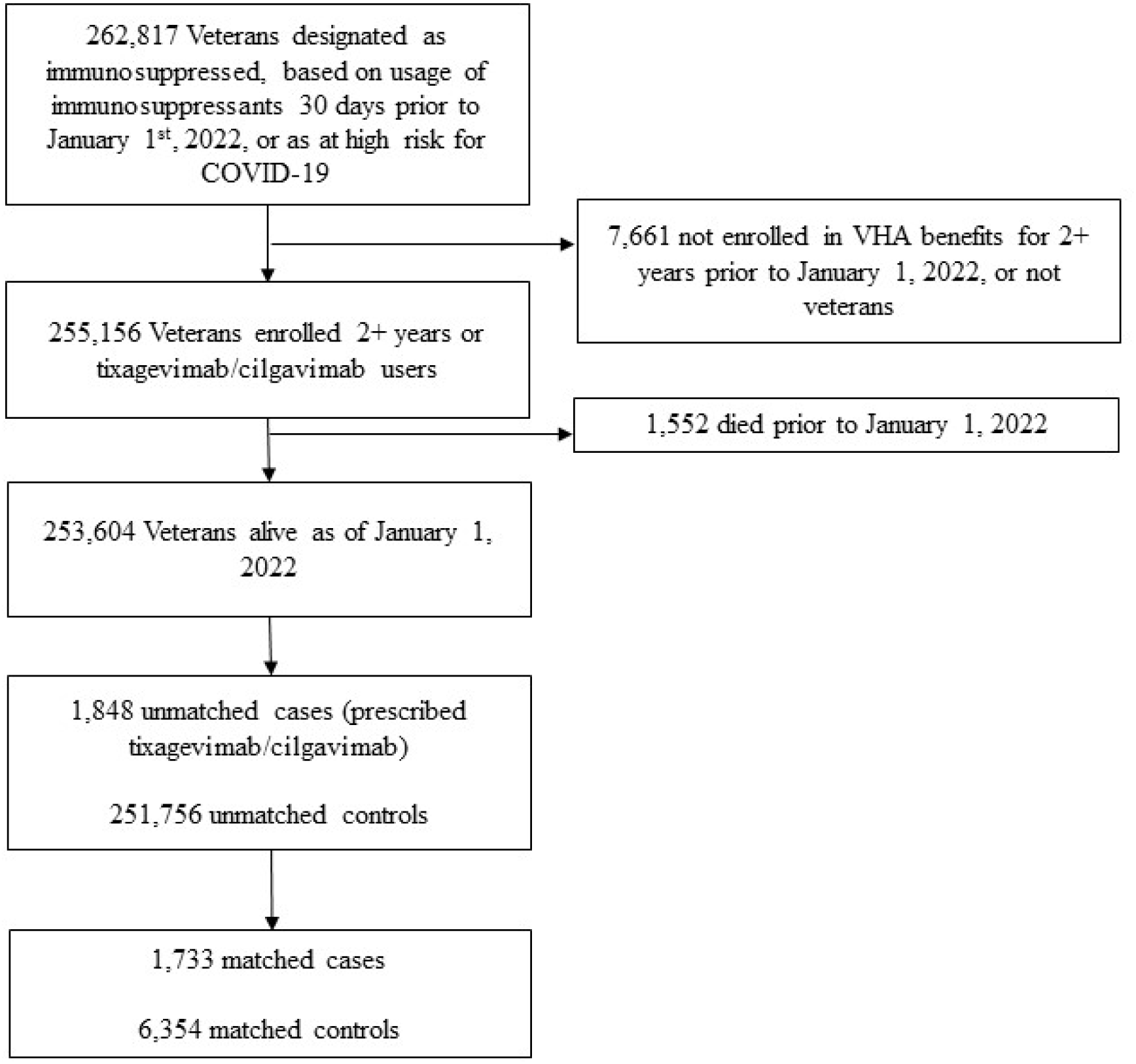
Selection of Patients.

To provide an overall picture of SARS-CoV-2 infection data for the study population during the extended study period, we displayed SARS-CoV-2 infection by calendar month from September 2021 to April 2022 with January 2022 as division when tixagevimab/cilgavimab first became available in the VA (**Figure 2**). Immunocompromised Veterans who received tixagevimab/cilgavimab (1,848) were shown next to the control group of 251,756 who did not receive tixagevimab/cilgavimab in categories of identified events: SARS-CoV-2 infection verified by positive PCR test, COVID-19-related hospitalization, all-cause mortality. During the last 4 months of 2021, before tixagevimab/cilgavimab became available at VA, the treatment group had an average incidence of SARS-CoV-2 infection or COVID-19-related hospitalization at 0.6% per month, while the control group had an average incidence of 0.8% per month. With the Omicron surge in January 2022, the first 4-month average in 2022 increased to 0.9% and 1.4% for the treatment and control groups, respectively. Although both groups experienced a surge, the treatment group was proportionally smaller than that of the control group (50% vs. 75% increase).

**Figure 2.**
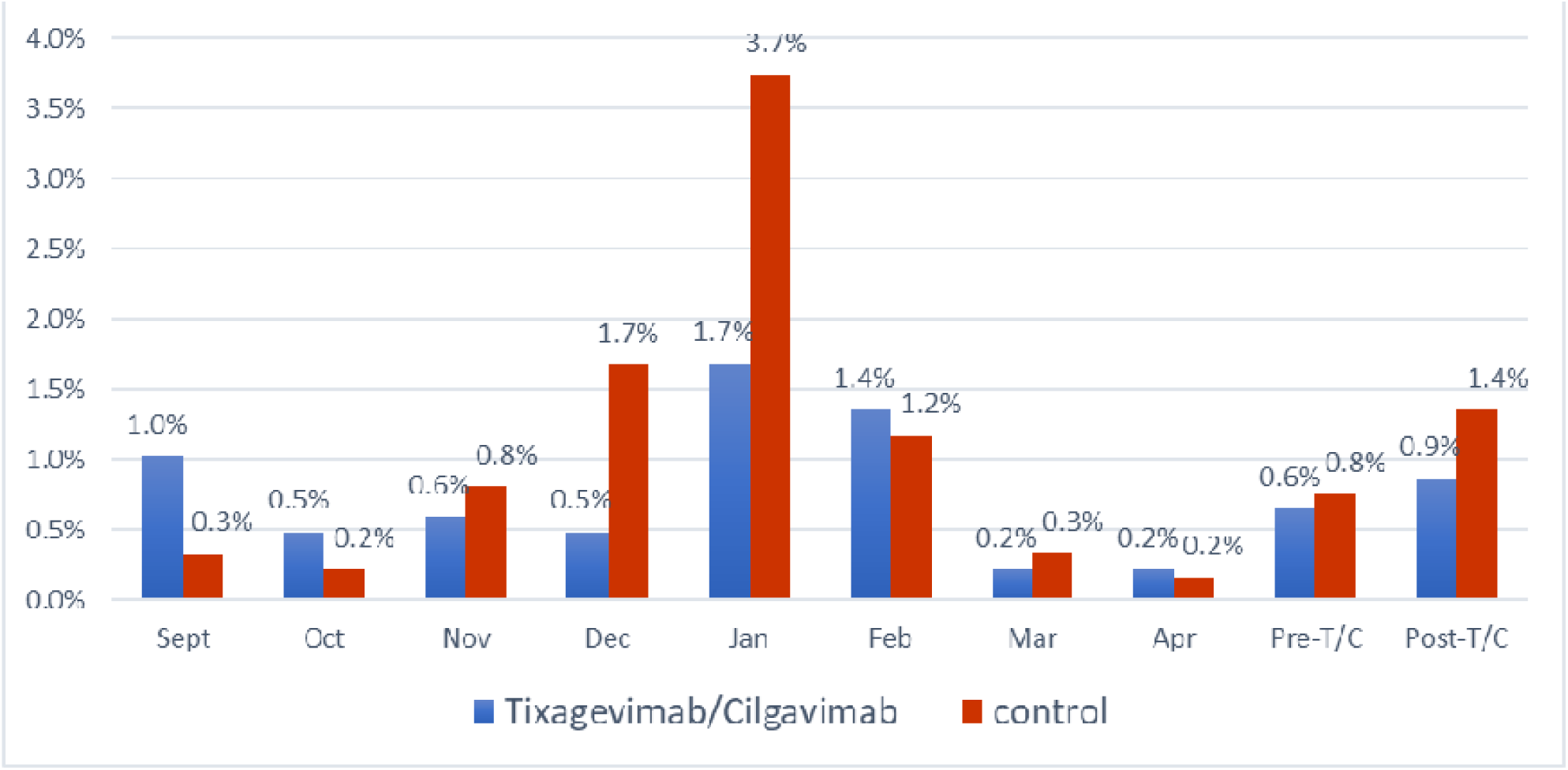
Rates of SARS-CoV-2 Infection and COVID-19 Hospitalization Before and After Availability of Tixagevimab/Cilgavimab at the VA (January 2022)

### Propensity Score Analysis

Estimated from propensity-score matched survival analyses, tixagevimab/cilgavimab recipients had a lower incidence of the composite of COVID-19 outcomes versus control patients overall (17/1733 [1.0%] vs 206/6354 [3.2%]; HR 0.31; 95%CI, 0.18-0.53). Results were similar within the study populations of EHR-confirmed immunocompromised (HR 0.32; 95%CI, 0.18-0.62), severely immunocompromised (HR 0.44; 95%CI, 0.21-0.93), and for Veterans aged 65 or older (HR 0.33; 95%CI, 0.18-0.61). The association in the overall cohort was similar across each of the individual COVID-19 outcomes, including test-confirmed SARS-CoV-2 infection (HR 0.34; 95%CI, 0.13-0.87), COVID-19 hospitalization (HR 0.13; 95%CI, 0.02-0.99), and all-cause mortality (HR 0.36; 95%CI, 0.18-0.73). (**Table 2, Figure 3**)

**Table 2.** Relative Effectiveness of Tixagevimab/Cilgavimab versus Untreated Controls using Propensity-Score Matched Analysis and Difference-in-Difference.

|  | Matched Controls<br>N=6,354 | Tixagevimab/cilgavimab recipients<br>N=1,733 | Propensity Score Survival Analysis | Difference in Difference^ Analysis |
| --- | --- | --- | --- | --- |
|  | Number of Events (%) | Number of Events (%) | Hazard Ratio (95% CI) | Incidence Rate Ratio (95% CI) |
| **Composite outcome (COVID-19 infection, COVID-19 hospitalization, and all-cause mortality)** |  |  |  |  |
| Overall Cohort | 206 (3.2%) | 17 (1.0%) | 0.31 (0.18-0.53) |  |
| Immunocompromised | 147 (3.5%) | 12 (1.0%) | 0.32 (0.18-0.62) |  |
| Severely Immunocompromised | 87 (3.7%) | 11 (1.4%) | 0.44 (0.21-0.93) |  |
| Not Immunocompromised* but at High Risk | 59 (2.8%) | (<1%)& | 0.27 (0.13-0.56) |  |
| **Individual Outcome (Overall Cohort)** |  |  |  |  |
| COVID-19 Infection | 69 (1%) | (<0.5%)& | 0.34 (0.13-0.87) | 0.32 (0.24-0.44) |
| COVID-19 related hospitalization | 38 (0.5%) | (<0.5%)& | 0.13 (0.02-0.99) | 0.10 (0.05-0.22) |
| All-cause Mortality | 99 (2%) | (<0.5%)& | 0.36 (0.18-0.73) |  |
| Falsification: Urinary Tract Infection | 127 (2%) | 36 (2%) | 1.05 (0.68-1.62) |  |

^ DiD analysis was not performed on outcomes involving mortality data because matched cohorts were all alive at index dates.
\* Electronic data regarding immunocompromised conditions or immunosuppressant use were found.
& Numbers not shown to protect patient information.

**Figure 3.**
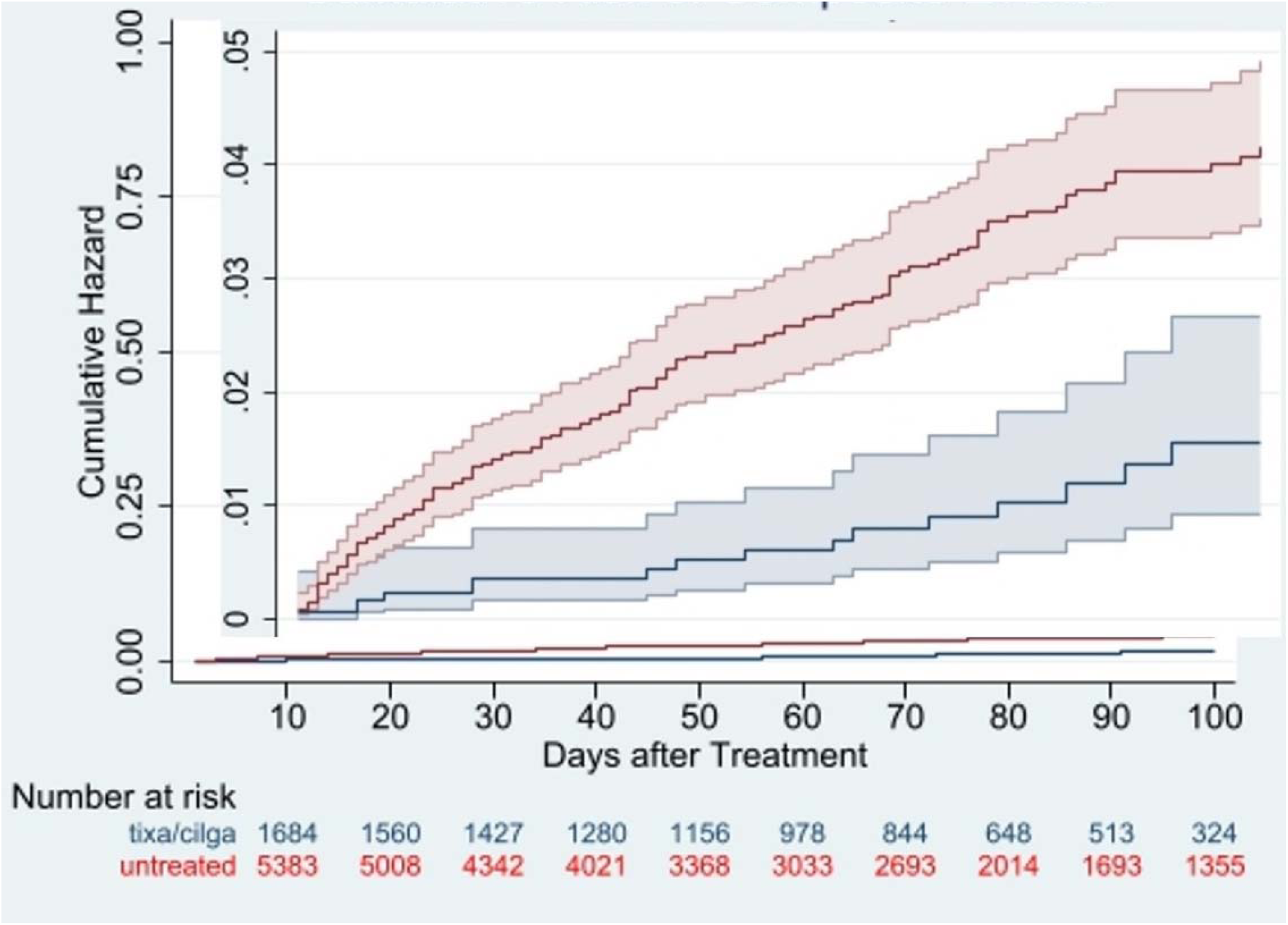
Cumulative Risk of Composite COVID-19 Outcomes for Tixagevimab-Cilgavimab Recipients Compared to Untreated Controls. Composite COVID-19 outcomes were SARS-CoV-2 infection, COVID-19 hospitalization, or all-cause mortality

Lastly, we were able to examine the impact of tixagevimab/cilgavimab with and without concomitant vaccination. Those fully vaccinated with at least 3 doses of any vaccine or 2 doses of Ad26.COV2, but without receiving tixagevimab/cilgavimab, had an incidence rate of 2.8% of COVID-19 infection or related hospitalization vs a rate of 3.7% among those neither vaccinated nor received tixagevimab/cilgavimab. Those not fully vaccinated but treated with tixagevimab/cilgavimab had an incidence rate of 1.35%. Most dramatically, those who were both fully vaccinated and received tixagevimab/cilgavimab had a rate of 0.85%, like the rate of a fully vaccinated and boosted non-immunocompromised adult. ^24^

### Sensitivity (DiD) Analysis

The interaction term between intervention (tixagevimab/cilgavimab vs control) and period (baseline and observation) was used to estimate the PERR-adjusted effectiveness using a Poisson regression model. The matched, PERR-adjusted effectiveness, as measure by incidence rate ratio was 0.32 (95% CI, 0.24-0.44%) against SARS-CoV-2 infection verified by a positive test, 0.10 (95% CI, 0.05-0.22) against COVID-19-related hospitalization, almost identical to the point estimates from propensity-scores matched survival analysis (**Table 2**). Because both actual and pseudo tixagevimab/cilgavimab use required the subjects to be alive, we were not able to perform PERR analysis on mortality, including the composite outcome.

### Falsification Analysis

Healthcare encounters with UTI as the primary discharge diagnosis were unlikely to be associated with tixagevimab/cilgavimab; therefore, served as a falsification test. One hundred sixty-three UTI visits were observed during the follow-up period. Propensity scores matched analysis demonstrated a similar effectiveness of tixagevimab/cilgavimab versus control against UTI (HR 1.05; 95% CI, 0.68-1.62) (**Table 2**). This lack of association between UTI and the treatment is reassuring that the protective effects associated with the treatment of tixagevimab/cilgavimab were unlikely due to bias or other major methodological flaws.

## DISCUSSION

In this retrospective cohort study using real-world data from patients across the VA healthcare system in the US, administration of tixagevimab/cilgavimab was associated with a significant reduction in the risk of SARS-CoV-2 infection, COVID-19 hospitalization, and all-cause mortality among patients who received tixagevimab/cilgavimab compared with controls. Our findings were consistent across two robust statistical approaches, including propensity score matching and DiD estimations. These findings were observed among immunocompromised, severely immunocompromised, and older patients, further supporting the EUA criteria for use of tixagevimab/cilgavimab in this population. Further, we found evidence of augmented protection against SARS-CoV-2 infections among fully vaccinated immunocompromised patients who received tixagevimab/cilgavimab, akin to that of the population of fully boosted adults who were not immunocompromised.^24^

To our knowledge, this is the first real-world evidence of tixagevimab/cilgavimab for prevention of COVID-19 and provides important insights regarding the patient population who have received tixagevimab/cilgavimab across VA healthcare system. Of note, the EUA encourages use of tixagevimab/cilgavimab primarily among fully vaccinated immunocompromised patients; however, none of the participants in the PROVENT trial were vaccinated. In comparison, **Error! Bookmark not defined**. nearly all (95%) of our study population received at least two doses of a COVID-19 mRNA vaccine or one dose of Ad26.COV2 before receiving tixagevimab/cilgavimab, and most (75%) were fully vaccinated. In PROVENT, only 11% of trial participants were immunocompromised, compared to at least 92% in the current analysis. Furthermore, patients aged 65 years and older accounted for a small proportion (24%) of patients included in the PROVENT trial, compared to 69% of tixagevimab-cilgavimab recipients in the current study. However, only a small proportion of eligible patients received treatment, indicating that enhanced education and outreach are paramount to ensure that more immunocompromised Veterans across the VA healthcare system receive this medication, specifically during COVID-19 surges.

In comparison to the PROVENT trial, the observation period of our analysis coincided with the Omicron BA.1 surge across the United States and provides important clinical data in this latest evolution of the pandemic. While tixagevimab/cilgavimab is maintained neutralization against the Delta variant of SARS-CoV-2, tixagevimab/cilgavimab was shown to have decreased neutralizing activity against the Omicron BA.1 variant, prompting the FDA’s revision of EUA to increase the initial dose tixagevimab/cilgavimab. Current data indicate that tixagevimab/cilgavimab maintains neutralization against Omicron BA.2 and BA.2.12.1, but this effect may be attenuated with BA.4 and BA.5.^25,26,27,28^

The present analysis supports the effectiveness of tixagevimab/cilgavimab in preventing SARS-CoV-2 infections caused by the Omicron variants, including predominantly BA.1 and the early BA.2 and BA.2.12.1 surge. Future longitudinal analyses will focus on newer Omicron variants. To help address this issue, the VA has launched several initiatives including VA SHIELD (<u>S</u>cience and <u>H</u>ealth Initiative to Combat Infectious and <u>E</u>merging <u>L</u>ife-threatening <u>D</u>iseases), a comprehensive biorepository of specimens from a cohort of affected Veterans with accompanying clinical data. As part of the future of VA SHIELD, clinical specimens will be collected prospectively from patients, which will help identify emerging strains as well as developing resistance in real world clinical settings. Obtaining this information rapidly will help public health officials, clinicians and researchers make important, timely decisions regarding diagnostics, prophylaxis, and therapeutics.

Our study had several notable strengths. We analyzed 1,486 patient-years of observation, making our study one of the largest ever conducted to assess tixagevimab/cilgavimab effectiveness while they were being distributed to combat a concurrent surge of the pandemic. The large sample allowed us to adjust for more potential confounding variables. Previous studies have shown that EHR data are more likely to be complete in capturing medical conditions and have a lower risk of up-coding. ^29,30^

Nevertheless, conventional analytical strategies such as stratification, matching (with or without propensity score), and multivariate regression analysis cannot adequately adjust for unobserved confounders. ^31,32,33^ These results were confirmed using two different statistical methods, including propensity-score matched models and DiD analysis. Propensity score matching of the intervention and comparison cohorts is an effective approach to control confounding. In addition, we employed an econometric technique – Difference-in-Difference (DiD) method - to adjust for bias from measured and unmeasured confounders and estimate the effectiveness of tixagevimab/cilgavimab in preventing COVID-19-related outcomes.

Finally, immortal time bias occurs when participants of a cohort study cannot experience the same outcome during a follow up period; if immortal time is misclassified or excluded during the analysis, the outcomes may be skewed. To account for immortal time bias in our analysis, we used a propensity-score matched survival analysis with Cox proportional hazards model to ensure controls were alive and COVID-free on the same day when their matched patients received tixagevimab/cilgavimab. This approach also ensured similar lengths of follow-up between the recipients and their matched controls.

### Limitations

There are some limitations to acknowledge. Firstly, VA data include only healthcare encounters occur in VA medical centers, so we could have missed some infections and hospitalizations that occurred outside VA, which could bias our results towards the null. Secondly, while the EUA criteria are intended for patients who are immunocompromised, a small proportion of patients (%) who received tixagevimab/cilgavimab were not immunocompromised based on our definition and it is possible that we misclassified these patients. Thirdly, the VA has a unique population (mostly male, older), and our results may not be generalizable to a larger population of patients that were not treated at the VA. ^34^ Fourthly, the 10th revision of the International Statistical Classification of Diseases and Related Health Problems (ICD-10) codes from claims data have been shown to inadequately capture comorbidity and functional status.^35^ Because only 289 (17%) of patients in our propensity-score matched tixagevimab/cilgavimab cohort received a single dose of 150 mg/150 mg tixagevimab/cilgavimab, we did not have the sufficient sample size to compare the original dosage of 150mg/150mg to the revised dosage of 300mg/300mg to assess the optimal dosing of tixagevimab/cilgavimab in the current analysis. Finally, we could not assess optimal timing of tixagevimab/cilgavimab in relation to COVID-19 vaccine administration, nor could we identify a target population who would be optimal to receive tixagevimab/cilgavimab.

### Conclusion

Using national real-world data from predominantly vaccinated, immunocompromised Veterans, we found that administration of tixagevimab/cilgavimab was associated with lower rates of SARS-CoV-2 infection, COVID-19 hospitalization, and all-cause mortality, compared with controls, during the Omicron surge. Our results suggest that tixagevimab/cilgavimab administration, in addition to vaccination, protects vulnerable patients from SARS-CoV-2 infection and severe COVID-19 in a contemporary phase of the pandemic. Ongoing real-world data will help to understand the effectiveness of tixagevimab/cilgavimab for pre-exposure prophylaxis over time and against emerging variants.

The contents of this article do not represent the views of the U.S. Department of Veterans Affairs or the U.S. Government. The views and opinions expressed in this article are those of the authors and do not necessarily reflect the official policy or position of the Food and Drug Administration, as well as any other agency of the U.S. Government. Assumptions made within and interpretations from the analysis do not necessarily reflect the position of any U.S. Government entity.

## Data Availability

Data in the present work will not be made available.

## Acknowledgements

We thank Dr. Hector Izurieta for his insight on observational study methods and guidance on real-world data.

## SUPPLEMENTARY APPENDIX

**Appendix I.** Immunosuppressants Used within 30 days before Index Date.

|  |  |  |  |  |
| --- | --- | --- | --- | --- |
| Abemaciclib | Chlorambucil | Glasdegib | Nilutamide | Tacrolimus |
| Abiraterone | Cladribine | Goserelin Acetate | Niraparib | Talazoparib |
| Acalabrutinib | Cobimetinib | Guselkumab | Obinutuzumab | Tamoxifen |
| Afatinib | Cortisone | Hydroxyurea | Ofatumumab | Tazemetostat |
| Alectinib | Crizotinib | Ibrutinib | Olaparib | Temozolomide |
| Alpelisib | Cyclophosphamide | Idelalisib | Omalizumab | Tepotinib |
| Anastrozole | Cyclosporine | Imatinib | Osimertinib | Thioguanine |
| Apalutamide | Dabrafenib | Infliximab | Palbociclib | Tivozanib |
| Apremilast | Darolutamide | Infliximab-Abda | Panobinostat | Topotecan |
| Asciminib | Dasatinib | Ivosidenib | Pazopanib | Toremifene |
| Atezolizumab | Degarelix | Ixazomib | Pemigatinib | Trametinib |
| Avapritinib | Dexamethasone | Ixekizumab | Ponatinib | Tretinoin |
| Axitinib | Duvelisib | Lapatinib | Pralsetinib | Triptorelin |
| Azacitidine | Enasidenib | Larotrectinib | Prednisolone | Tucatinib |
| Azathioprine | Encorafenib | Letrozole | Prednisone | Umbralisib |
| Belumosudil | Entrectinib | Leuprolide | Procarbazine | Ustekinumab |
| Belzutifan | Enzalutamide | Lomustine | Regorafenib | Vandetanib |
| Bevacizumab | Erdafitinib | Lorlatinib | Relugolix | Vemurafenib |
| Bexarotene | Erlotinib | Lurbinectedin | Ribociclib | Venetoclax |
| Bicalutamide | Etanercept | Melphalan | Rilonacept | Vismodegib |
| Binimetinib | Etoposide | Mercaptopurine | Ripretinib | Vorinostat |
| Bosutinib | Everolimus | Methotrexate | Rucaparib | Zanubrutinib |
| Brigatinib | Exemestane | Methylprednisolone | Secukinumab |  |
| Budesonide | Fludrocortisone | Midostaurin | Selinexor |  |
| Busulfan | Fluorouracil | Mitomycin | Selpercatinib |  |
| Cabozantinib | Flutamide | Mitotane | Sirolimus |  |
| Canakinumab | Fulvestrant | Mycophenolate Mofetil | Sonidegib |  |
| Capecitabine | Gefitinib | Mycophenolic Acid | Sorafenib |  |
| Capmatinib | Gemcitabine | Neratinib | Sotorasib |  |
| Ceritinib | Gilteritinib | Nilotinib | Sunitinib |  |

**Appendix II.** Definitions of Patient Characteristics including Immunocompromised Conditions.

| Variable | Values | Definition | Timing |
| --- | --- | --- | --- |
| Age | Integer | Age as 31 Dec 2021 | Table 1: Age as of 31 Dec 2021 |
| Sex | Female/male | As defined in VHA data | Most recent available |
| Race/ethnicity | Non-Hispanic Black<br>Hispanic any race<br>Non-Hispanic White<br>Other | Black: non-Hispanic black<br>Hispanic: Hispanic any race<br>White: non-Hispanic white<br>Other: non-Hispanic other race, missing race, declined to state, unknown | Most recent available |
| Rurality | Highly Rural<br>Rural<br>Urban | VHA defined based on the Rural-Urban Community Area (RUCA) system [https://www.ruralhealth.va.gov/rural-definition.asp] | Most recent available |
| VHA defined priority group | 1, 2, 3, 4, 5, 6, 7, 8 | VA defined based on factors including military service history, disability rating and income level to identify Veterans to determine enrolment priority; 1 is the highest priority [https://www.va.gov/health-care/eligibility/priority-groups/] | Most recent available |
| Nursing home use | 0/1 | 1 = Any nursing home or long-term care indicated on an inpatient admission or place of disposition | During 2 years before Index date (from 01 Jan 2019 through Index date) |
| DCCI | Numeric | Using ICD-10 codes from any inpatient or outpatient record, computed Deyo-Charlson Comorbidity Index. Coding algorithms for defining comorbidities in ICD-9-CM and ICD-10 administrative data. Medical Care 2005; 43(11):1130-1139. DOI: 10.1097/01.mlr.0000182534.19832.83] | During 2 years before Index date |
| BMI | Normal<br>Overweight/obese<br>Missing | Normal: BMI less than 26<br>Overweight/obese: BMI greater than or equal to 26 | Most recent available from 01 Jan 2018 through 30Apr2022 |
| Comorbidities |  | All comorbidities listed below have 1 inpatient or 2 outpatient records with the corresponding ICD-10 code | During 2 years before Index date. |
| Asthma | 0/1 | ICD-10 code J45*<br>ICD-10 code J44.9<br>ICD-10 code J67.8 |  |
| Cancer | 0/1 | Charlson condition definitions using ICD-10 codes |  |
| Cancer metastatic | 0/1 | Charlson condition definitions using ICD-10 codes |  |
| Coronary artery disease | 0/1 | ICD-10 code I 25* |  |
| Congestive heart failure | 0/1 | Charlson condition definitions using ICD-10 codes |  |
| Chronic kidney disease | 0/1 | ICD-10 code N18* |  |
| Chronic obstructive pulmonary disease | 0/1 | Charlson condition definitions using ICD-10 codes |  |
| Cardiovascular disease | 0/1 | Charlson condition definitions using ICD-10 codes |  |
| Dementia | 0/1 | Charlson condition definitions using ICD-10 codes |  |
| Diabetes mellitus with complications | 0/1 | Charlson condition definitions using ICD-10 codes |  |
| Diabetes mellitus without complications | 0/1 | Charlson condition definitions using ICD-10 codes |  |
| Dyslipidemia | 0/1 | ICD-10 code E78.5 |  |
| HIV | 0/1 | Charlson condition definitions using ICD-10 codes |  |
| Hypertension | 0/1 | ICD-10 code H35.03*<br>ICD-10 code I10*<br>ICD-10 code I11*<br>ICD-10 code I12*<br>ICD-10 code I13*<br>ICD-10 code I15*<br>ICD-10 code I16<br>ICD-10 code I67.4 |  |
| Liver disease, mild | 0/1 | Charlson condition definitions using ICD-10 codes |  |
| Liver disease, severe | 0/1 | Charlson condition definitions using ICD-10 codes |  |
| Myocardial infarction (history) | 0/1 | Charlson condition definitions using ICD-10 codes |  |
| Para/hemiplegia | 0/1 | Charlson condition definitions using ICD-10 codes |  |
| Peptic ulcer disease | 0/1 | Charlson condition definitions using ICD-10 codes |  |
| Peripheral vascular disease | 0/1 | Charlson condition definitions using ICD-10 codes |  |
| Rheumatoid arthritis | 0/1 | Charlson condition definitions using ICD-10 codes |  |
| Renal disease | 0/1 | Charlson condition definitions using ICD-10 codes |  |
| Atrial fibrillation | 0/1 | ICD-10 code I48.0<br>ICD-10 code I48.11<br>ICD-10 code I48.19<br>ICD-10 code I48.21<br>ICD-10 code I48.91 |  |
| Anaphylaxis (history) | 0/1 | ICD-10 code Z87.892 |  |
| Arthritis | 0/1 | ICD-10 code M15* through M19* |  |
| Bleeding diathesis | 0/1 | ICD-10 code D69.9 |  |
| Bronchiectasis | 0/1 | ICD-10 code J47* |  |
| Depression | 0/1 | ICD-10 code F33* |  |
| Down Syndrome | 0/1 | ICD-10 code Q90* |  |
| Embolism (history) | 0/1 | ICD-10 code I82*<br>ICD-10 code I74*<br>ICD-10 code I26* |  |
| Falls (history) | 0/1 | ICD-10 code W19* |  |
| Gout | 0/1 | ICD-10 code M1A* |  |
| Hepatitis B | 0/1 | ICD-10 code B19.1* |  |
| Hepatitis C | 0/1 | ICD-10 code B19.2* |  |
| Hyperlipidemia | 0/1 | ICD-10 code E78.2<br>ICD-10 code E78.41<br>ICD-10 code E78.49<br>ICD-10 code E78.5 |  |
| Hypersensitivity<br>pneumonitis | 0/1 | ICD-10 code J67* |  |
| Interstitial lung disease | 0/1 | ICD-10 code J80*<br>ICD-10 code J81*<br>ICD-10 code J82*<br>ICD-10 code J84* |  |
| Impaired mobility | 0/1 | ICD-10 code Z74.01<br>ICD-10 code Z74.09 |  |
| Musculoskeletal disorder | 0/1 | ICD-10 code M00* through<br>M99* |  |
| Mycoses | 0/1 | ICD-10 code B35* through<br>B49* |  |
| Obesity | 0/1 | ICD-10 code E66* |  |
| Paranoia | 0/1 | ICD-10 code F22*<br>ICD-10 code F60* |  |
| Parkinson's | 0/1 | ICD-10 code G20* |  |
| Pregnancy (history) | 0/1 | ICD-10 code Z33* |  |
| Drug-induced anaphylaxis | 0/1 | ICD-10 code Z88* |  |
| Sickle cell disease | 0/1 | ICD-10 code D57* |  |
| Solid organ transplant<br>recipient | 0/1 | ICD-10 code Z94* |  |
| Stroke / transient<br>ischemic attack | 0/1 | ICD-10 code I63*<br>ICD-10 code G45* |  |
| Urinary tract infection | 0/1 | ICD-10 code N39.0 |  |
| Immunocompromised<br>Conditions | 0/1 | ICD-10 code B20*<br>ICD-10 code B59*<br>ICD-10 code B97.3*<br>ICD-10 code D47.Z1*<br>ICD-10 code D70*<br>ICD-10 code D71*<br>ICD-10 code D72*<br>ICD-10 code D73*<br>ICD-10 code D76* |  |
|  |  | ICD-10 code D80*<br>ICD-10 code D81*<br>ICD-10 code D82*<br>ICD-10 code D83*<br>ICD-10 code D84*<br>ICD-10 code D89*<br>ICD-10 code M05*<br>ICD-10 code M06*<br>ICD-10 code M07*<br>ICD-10 code M08*<br>ICD-10 code M30*<br>ICD-10 code M31*<br>ICD-10 code M32*<br>ICD-10 code M33*<br>ICD-10 code M34*<br>ICD-10 code M35.0*<br>ICD-10 code M35.9*<br>ICD-10 code Q89.0*<br>ICD-10 code T45.1X1<br>ICD-10 code Z21*<br>ICD-10 code Z48.2*<br>ICD-10 code Z51.0*<br>ICD-10 code Z51.1*<br>ICD-10 code Z94*<br>ICD-10 code R76*<br>ICD-10 code Z79.5<br>ICD-10 code C00-C26<br>ICD-10 code C30-C34<br>ICD-10 code C37-C41<br>ICD-10 code C43-C58<br>ICD-10 code C60-C88<br>ICD-10 code C4A<br>ICD-10 code C7A<br>ICD-10 code C7B<br>ICD-10 code C90-C96<br>ICD-10 code D03<br>ICD-10 code D46<br>ICD-10 code Z85<br>[[https://www.cdc.gov/vaccines/covid-19/downloads/us-flu-vaccine-effectiveness-network-protocol-508.pdf]](https://www.cdc.gov/vaccines/covid-19/downloads/us-flu-vaccine-effectiveness-network-protocol-508.pdf) |  |
| Hematological malignancy | 0/1 | ICD-10 codes 81.70, 81.79, 81.90<br>ICD-10 codes 82.90<br>ICD-10 codes 84.10, 84.40<br>85.80<br>ICD-10 code C88.0<br>ICD-10 code C90.00<br>ICD-10 codes C91.00, 91.01, 91.10, 91.11, 91.40<br>ICD-10 codes C92.00, 92.01, 92.10, 92.11, 92.30<br>96.2<br>ICD-10 code D47.2 |  |
| GvHd | 0/1 | ICD-10 code : D89.81 |  |
| Vaccination status | None<br>Full<br>Booster | None: until first vaccination date<br>Full: from 14 days after the second vaccination date<br>Booster: from 14 days after the third vaccination date | Determined at date of treatment |
| Vaccine manufacturer | None<br>Moderna<br>Pfizer<br>Janssen<br>AstraZeneca | None: no vaccination recorded<br>Moderna: CPT codes 91301, 0011A, 0012A, 0013A, 91306, 0064A or vaccination name<br>Pfizer: CPT codes 91300, 0001A, 0002A, 0003A, 0051A, 0052A, 0053A, 0054A or vaccination name<br>Janssen: 91303, 0031A, 0034A, or vaccine name<br>AstraZeneca: 91302, 0021A, 0022A, or vaccine name | Determined at date of first vaccination |
| CAN Score |  | Score to identify patients at highest risk of hospitalization and mortality. As indicated in VHA data. If missing, the score is assumed to be zero. [https://www.va.gov/HEALTHCAREEXCELLENCE/about/organization/examples/care-assessment-needs.asp] | Most recent CAN score during 2 years before and up to end of study period 01Jan2018 through 30Apr2022 |
| Mortality-1 year | Percentage of probability (part of CAN score) | Measurement of risk of mortality in next year |  |
| Mortality-90 days | Percentage of probability (part of CAN score) | Measurement of risk of mortality in the next 90 days |  |
| Event-1 year | Percentage of probability (part of CAN score) | Measurement of risk of any visit or event in the next year |  |
| Event-90 days | Percentage of probability (part of CAN score) | Measurement of risk of any visit or event in the next 90 days |  |
| Hospitalization-1 year | Percentage of probability (part of CAN score) | Measurement of risk of a hospitalization in the next year |  |
| Hospitalization-90 days | Percentage of probability (part of CAN score) | Measurement of risk of a hospitalization in the next 90 days |  |
| Number of encounters | Integer | Count of VHA related encounters (inpatient or outpatient, including telemedicine) | Look back period is 01Jan2019 to Index date |
| Number of shots | Integer 0-2 | 0 = not vaccinated<br>1 = partially vaccinated with a 2-dose mRNA<br>2 = vaccinated with 2-dose series of mRNA or 1 Janssen | Vaccination before treatment |
| Covid-19 before the Evusheld era | 0/1 | Positive SARS-CoV-2 lab test at VA before Index date | Positive SARS-CoV-2 lab test at VA before Index date |
| Other control | 0/1 | Patients who are at high risk of COVID-related morbidity and mortality (Obesity, liver disease, CHF, CKD, COPD, adverse event from a vaccination) but for whom no records of either immunocompromised conditions or use of immunosuppressants were found. | Looked for event 01Jan2019 through Index date |
| Inpatient Admission for Covid | 0/1 | Having an inpatient admission with both an admission and discharge diagnosis with COVID-19 and within 30 days of a positive SARS-CoV-2 lab test. ICD10-CM codes: U07.1, J12.82, M35.81, Z20.822, M35.89 |  |

**Appendix III.**
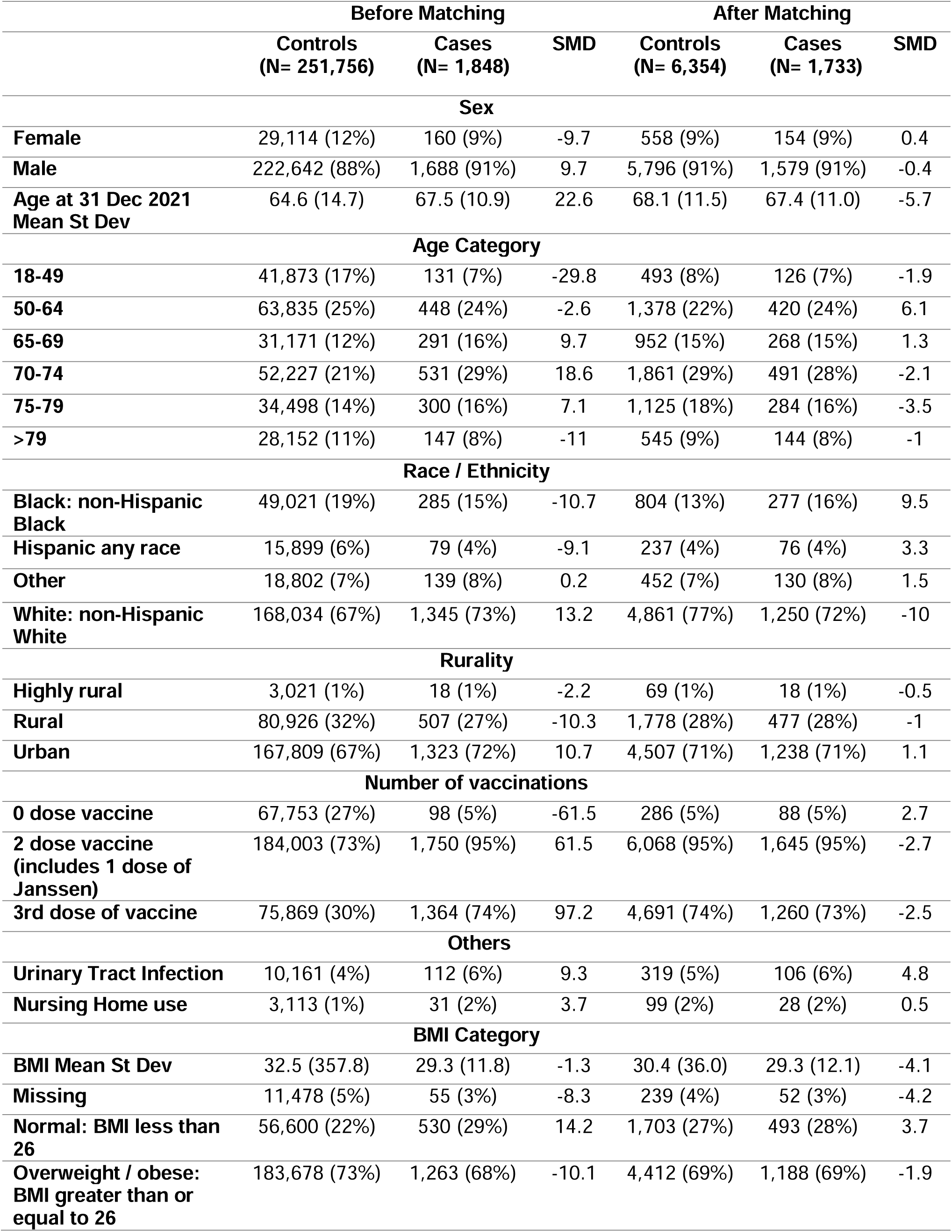
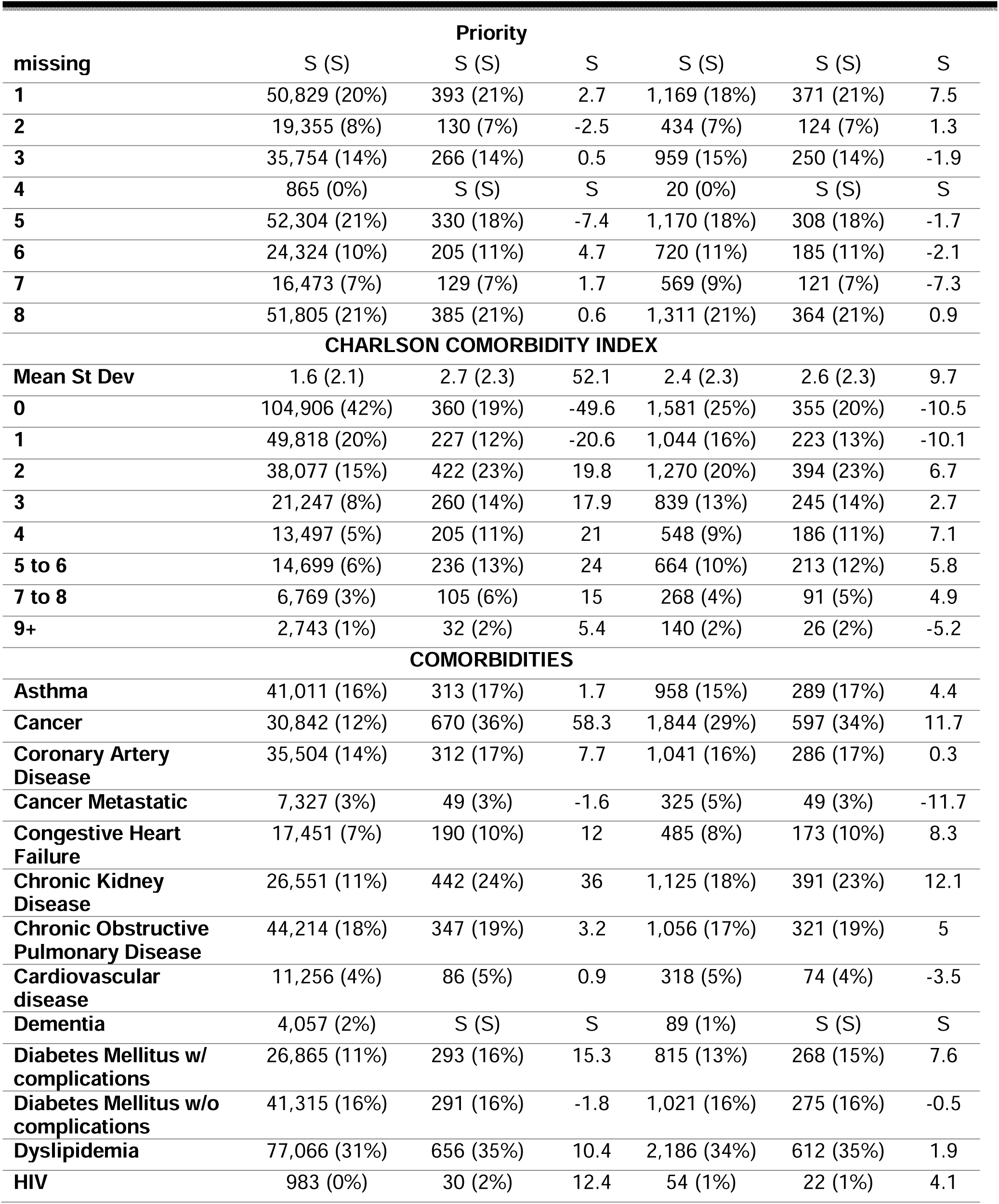
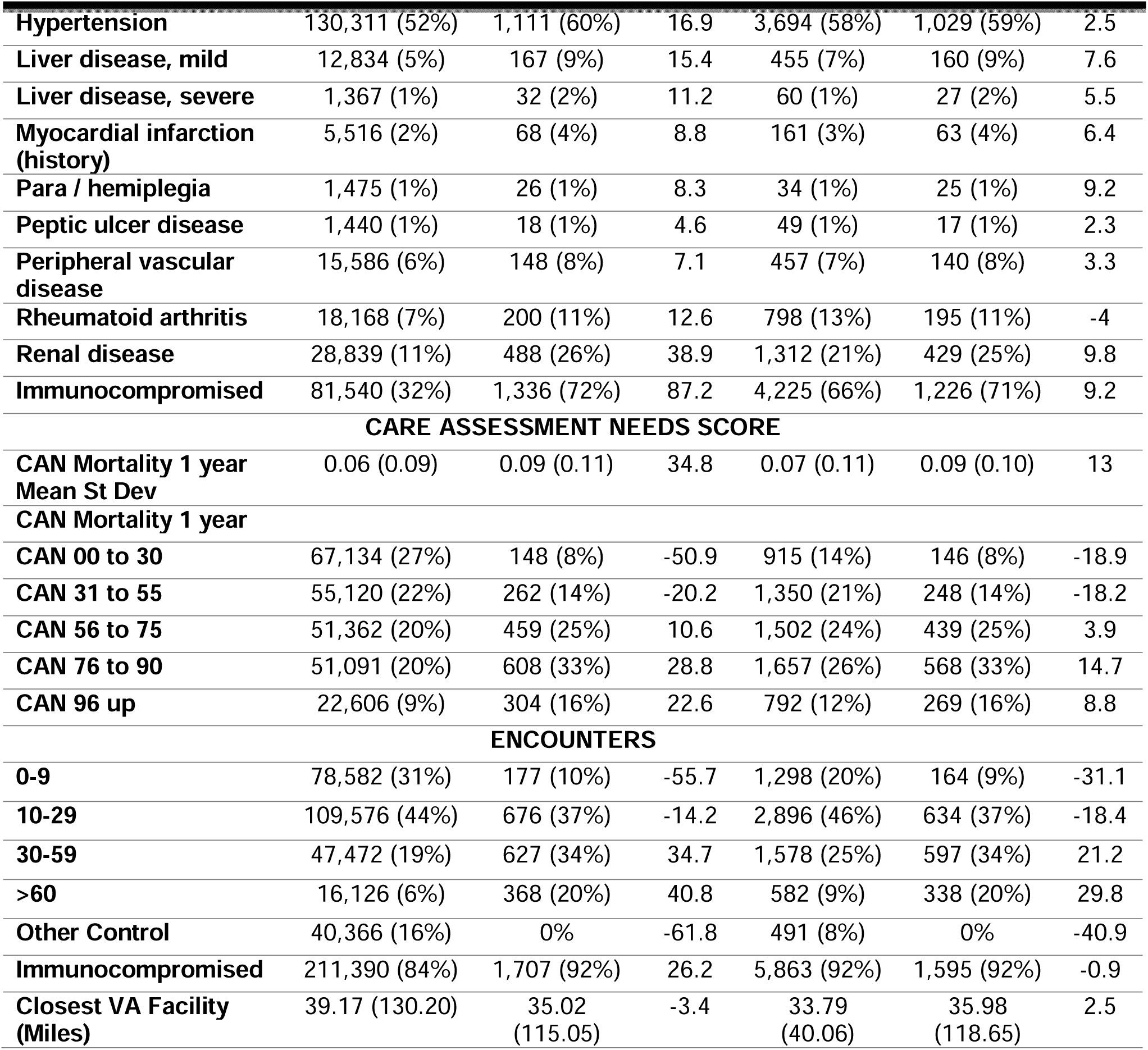
All Baseline Characteristics.

